# From Volume to Value: Comparative value, quality, and cost between three treatment modalities for early childhood caries

**DOI:** 10.1101/2022.02.04.22270476

**Authors:** David Okuji, Ashiana Jivraj, Yinxiang Wu

**Affiliations:** NYU Langone Dental Medicine, Hansjörg Wyss Department of Plastic Surgery, Division of Dental Medicine, NYU Grossman School of Medicine, 5800 Third Avenue, 3^rd^ Floor, Brooklyn, NY 11220; Harvard School of Dental Medicine, 188 Longwood Avenue, Boston, MA 02115; NYU School of Medicine-Division of Biostatistics, Department of Population Health, 180 Madison Avenue, Fifth Floor, New York, NY 10016

**Author notes:** **Contribution:** Concept, design, definition of intellectual content, literature search, clinical studies, data acquisition, manuscript preparation, manuscript editing, manuscript review. **Contribution:** Literature search, manuscript preparation, manuscript editing, manuscript review. **Contribution:** Literature search, data analysis, statistical analysis, manuscript preparation, manuscript editing, manuscript review. **Corresponding Author:** David Okuji, DDS, MBA, MS, NYU Langone Hospitals, 5800 Third Avenue, 3^rd^ Floor, Brooklyn, NY 11220, USA, Facsimile.

**Keywords:** child, dental-caries, treatment, quality, cost, surgical, disease-management, general-anesthesia

## Abstract

**Context:** In recent decades there has been a shift from a reimbursement system based upon treatment “volume” to “value.”

**Aims:** To compare value, quality, and cost of three treatment modalities for early childhood caries: surgical care with and without general anesthesia, and non-surgical care with disease management.

**Settings and Design:** Through an analytical, observational, cross-sectional design, data was collected from 487 pediatric subjects treated at health centers in seven states. Nearly 95% of the subjects were beneficiaries of the United States government Medicaid health insurance program, which supports families with low socioeconomic status.

**Methods and Material:** Quality was measured with a Parental-Reported Symptom and Service Quality questionnaire, adapted from the validated Early Child Oral Health Impact Scale, Child-Oral Impacts on Daily Performance, and Dental Consumer Assessments of Healthcare Providers and Systems surveys. Cost was measured by the labor cost for clinicians and adjusted by total relative value units for all treatment procedures. Value was calculated as the quotient of the weighted quality score and the adjusted cost, multiplied by a coefficient.

**Statistical Analysis:** Value comparisons were analyzed using chi-square, ANOVA, and likelihood ratio tests.

**Results:** Children treated with non-surgical, disease management had significantly higher parental-perceived quality, lower cost, and higher value when compared to the two surgical treatment modalities.

**Conclusions:** Dentists should consider that non-surgical treatment yields higher “value” than surgical treatments for the management of early childhood caries. Payers and consumers would benefit by comparing treatment “value” when choosing institutions and providers for the management of early childhood caries.

## Introduction

Early Childhood Caries (**ECC**), generally described as tooth decay among children younger than 6 years of age, is highly prevalent with 23 percent of United States (**U.S**.) pre-school children experiencing dental caries, and costly with $892 million in total U.S. dental expenditures in 2012.^[1,2}^ In 2015-2016, the prevalence of ECC dropped slightly to 21% nationally and disproportionately affected ethnic minorities and the poor.^[3]^

The Affordable Care Act (**ACA**) of 2010 addresses access to care and reducing the burden of ECC by including pediatric oral health as an essential health benefit codified through U.S. federal law.^[4]^ As well, the ACA intends to move health care institutions and providers to improve care for individuals, improve the health of the population, and reduce the cost of care through the “Triple Aim” objectives of “better care, better health, at lower cost” which are advocated by the Institute of Medicine (now named the National Academy of Medicine) and the Institute for Healthcare Improvement.^[5]^ The slight decrease in ECC prevalence between 2012 and 2016 could be attested to the ACA impacts of Medicaid and Children’s Health Insurance Program (**CHIP**) expansions, which have increased health care access to children and adolescents by over 50%.^[6]^

In the 2016 textbook, “Redefining Health Care: Creating Value-Based Competition on Results,” Professors Michael Porter and Elizabeth Teisberg at the Harvard Business School describe a detailed value-based health care model for meeting the “Triple Aim” objectives by focusing on specific medical conditions over cycles of care, measuring the patient-perceived quality of care, and measuring the direct cost of care expended by health care delivery institutions and providers.^[7]^ Specifically, Porter and Teisberg defined “Value” of care as the quotient of “Quality” and “Cost” (V = Q/C) and they further challenged the U.S. health care system, government, institutions, payers, and providers of health care to align structures, processes, and outcomes in order to increase value to patients.^[8]^

Current reimbursement mechanisms for U.S. health care facilities and providers are still predominantly driven by volume. As examples for how institutions and providers are paid; in a fee-for-service scheme, reimbursement is driven by the volume of procedures; in a prospective payment scheme, by the volume of patient visits; and in a capitation scheme, on a per member per month basis and the volume of enrolled members.^[9]^ The fee-for-service and prospective payment schemes motivate institutions to increase the volume of procedures or visits, respectively. The capitation scheme motivates institutions to maximize the volume of enrolled members while minimizing the volume of visits and procedures, without requiring the institutions and providers to address the quality of outcomes.^[10]^ Thus, the ACA mandate is to move from a system of care driven by “volume” to a system driven by “value”, by measuring the level of the quality of health outcomes relative to the cost of care.^[11]^

Traditionally ECC is treated surgically in the out-patient clinic or in the operating room under general anesthesia, especially in cases of uncooperative children.^[12]^ However, managing the etiology of caries and stopping the spread with disease management protocols has also become an important method of treating this rampant disease.^[13]^ Through the lens of the Porter-Teisberg model, comparative value between these treatment modalities permit ranking of their effectiveness for better outcomes. In addition, creating an easily applied value-based model can help clinicians decide which treatment options are more effective for their pediatric patients. The value model may also allow for employers, who make the bulk of insurance-buying decisions; and payers for care, such as Medicaid and commercial insurance companies; to identify dentists who provide the highest value for care. As well, the model may help parents, who do not have dental insurance benefits, to compare and select dentists on the basis of higher value.

The purpose of this study is to utilize a simplified adaptation of the Porter-Teisberg model to compare quality, cost, and value between three ECC treatment modalities; surgical care under general anesthesia (**GA**), surgical care without GA, and non-surgical care with disease management; with the hypothesis that non-surgical care with disease management treatment would yield higher value than the two surgical modalities.

## Methods

### Overview

An analytical, observational, cross-sectional design was used to describe and analyze the results of a methodology to measure value, quality, and cost for the management of ECC. The study was approved by the NYU Grossman School of Medicine Institutional Review Board under protocol number 16-00331.

### Study Population

The study population was a stratified, convenience sample of 487 subjects recruited at seven federally qualified health centers (**FQHCs**) located in Arizona, Florida, Hawaii, Maryland, Massachusetts, New York, and Tennessee. The subjects met the inclusion criteria and were stratified by treatment modality at the initial treatment visit. The post-hoc power analysis was calculated at 99.4%, based upon the study’s sample size, treatment size effects, and alpha at 0.05.

### Definitions of Data Outcomes

This study defined “value,” “quality,” and “cost,” respectively, as the value of care provided to the pediatric patient; the quality of care perceived by the patient’s parent; and the cost of care as the total direct labor cost expended by the health care institution to provide the care to the patient, divided by cumulative relative value units for the procedures performed.

Quality was measured with a 28-question Parental-Reported Symptom and Service quality Questionnaire (**PRSSQ**), a post-treatment survey administered to the parents of the ECC treated children, one month after the final treatment visit. The PRSSQ was adapted and compiled from three validated and parental-reported clinical and satisfaction surveys consisting of 13-questions from the Early Child Oral Health Impact Scale (**ECOHIS**), 6-questions from the Child-Oral Impacts on Daily Performance (**C-OIDP**), and 9-questions from the Dental Consumer Assessments of Healthcare Providers and Systems (**D-CAHPS**).^[14–16]^ The ECOHIS assesses the effects of the child’s oral health problems on not only the child, but also on the child’s parent or caregiver.^[14]^ The C-OIDP assesses oral impacts on the following daily performances: eating, speaking, cleaning teeth, smiling, emotional stability, relaxing, doing schoolwork, and social contact.^[16]^ The D-CAHPS assesses the dental care experiences and satisfaction level of consumer-caregivers.^[15]^ Due to the disparities with the number of PRSSQ questions ascribed to the ECOHIS (13-questions), C-OIDP (6-questions), and D-CAHPS (9-questions); the average PRSSQ quality score was calculated as the weighted average score for the combined three validated surveys.

To simplify the Porter-Teisberg determination of cost, total direct labor cost was utilized as a proxy for total direct cost since direct labor cost contributes to nearly 60% of operational expenses for a pediatric dental practice.^[17]^ Accountants define “total direct cost” as the cost of materials and supplies, in addition to labor. This study defined “direct labor cost” as the total labor cost for dentists, dental-residents, anesthesiologists, nurse-anesthetists, nurses, operating room technicians, and dental assistants required to complete the treatment for each child. Direct labor cost was measured, and adjusted for the effects of the types and number of total procedures, by calculating the total duration of treatment time for the total number of visits, multiplying by the labor rate for the aggregate of each clinical staff member involved with the child’s treatment, and dividing by the total Relative Value Units (**RVUs**) for the types and total number of procedures performed for all treatment visits.^[18]^ In this way, the RVU-adjusted cost provided a basis for equal comparison among the three treatment modalities such that the Value (**V**) was calculated as quality (PRSSQ score), divided by RVU-adjusted cost (**C**), times a coefficient of 100, with the multiplier product resulting in a value numeric which is greater than zero.

### Data Collection

Data was collected between December 1, 2015 and October 3, 2016 from 487 subjects at FQHCs and hospitals affiliated with the NYU Langone Hospitals Dental Post-Doctoral Residency Programs located in Arizona, Florida, Hawaii, Maryland, Massachusetts, New York, and Tennessee. The FQHCs and hospitals in the seven states serve communities with underserved and multi-race/ethnic compositions. The subjects were recruited from the parents/legal caregivers of pediatric patients, with a diagnosis of ECC, who underwent the informed consent process and opted for surgical treatment under GA, surgical treatment without GA, or non-surgical treatment. Surgical care with general anesthesia involved full-mouth dental rehabilitation in one visit under GA. Surgical care without GA involved full-or partial-mouth treatment with local anesthesia in one or more treatment visits with or without the use of nitrous oxide analgesia. Non-surgical disease management care involved treatment with topical fluoride varnish and silver diamine fluoride (**SDF**) applications, interim therapeutic restorations, motivational interviewing, frequent follow-up treatment visits, and active surveillance visits.

Pediatric patients were included if they were registered at an NYU Langone Hospitals affiliated FQHC, enrolled in the study at age 6 years or younger, diagnosed with ECC as defined by the American Academy of Pediatric Dentistry (**AAPD**), had a full complement of 20 primary teeth, and whose parents were able to consent for treatment in Spanish or English language. The patients completed an oral examination with a treatment plan; were classified as an American Society of Anesthesiologists (**ASA**) Physical Status Classification 1, 2, or 3; and had the treatment modality categorized as either surgical care with general anesthesia, surgical care without general anesthesia, or non-surgical disease management care. Patient treatment had to be completed in five or fewer visits.

During the treatment visits, the time of ingress for the subject entering the treatment room and the time of egress for the subject leaving the treatment room were recorded. As well, the number and types of clinical personnel assigned to care for the patient’s treatment were recorded. The duration of treatment, in hours, multiplied by the aggregated hourly gross wage rate for each of the clinical personnel caring for the patient’s treatment yielded the gross, direct labor cost for the facility to provide the treatment.

The post-treatment PRSSQ survey, measured the parental-perceived quality of care for their children, was administered by telephone or in-person one-month after the treatment was completed.

### Statistical Methods

Counts and percentages were calculated for categorical variables. Means and standard deviations were calculated for continuous variables. Comparisons of sample demographic characteristics; survey responses; and quality, cost, and value of care by treatment modalities (surgical with GA, surgical without GA, and non-surgical) were performed by the chi-square test for categorical variables and the ANOVA test (or the Kruskal-Wallis test, as appropriate) for continuous variables. Natural Log-transformed (**ln**) value scores, which have an approximately normal distribution, were also included in the comparison.

The distribution of three treatment modalities was presented across states. Since not all treatment modalities were observed in every state, only the data from Maryland and New York states were used to compare the value of all three treatment modalities. The data from all states except Hawaii were used for the comparison between treatment modality surgical with GA and surgical without GA. Linear mixed-effect models were fitted to estimate the effects of treatment modalities on the ln-transformed value, with adjustments for patient age, gender, race/ethnicity, and ASA status. Because the data had multiple levels of nesting, which included patient (level 1), dentist (level 2), clinic facility (level 3), and state (level 4), the likelihood ratio test was used to choose how many levels of nesting were needed in the model.

All analyses were performed in R version 3.5.0 for Windows (The R Foundation for Statistical Computing, Vienna, Austria.), using the package ‘nlme’ for linear mixed-effect models.^[19,20]^

## Results

The analytic sample included 487 pediatric patients. Overall, 49.9% of the patients were male, 56.2% were Hispanic, and the mean (standard deviation (**SD**)) age was 3.99 (1.22) [Table 1]. The majority (87.5%) of the sample were healthy with ASA level 1. Table 4 shows the distributions of treatment modalities by states, indicating that only the training locations in Maryland and New York collected data for all three treatment modalities. The non-surgical modality, which includes the use of SDF as a caries-arresting agent, was not utilized at the other five training locations because SDF was not commercially available in the U.S. until 2015 and these five locations had not yet implemented the use of SDF during the data collection period.

**Table 1.** Summary of sample characteristics by treatment modalities.

|  | Overall | Surgical with GA | Surgical without GA | Non-Surgical | p |
| --- | --- | --- | --- | --- | --- |
| N (%) | 487 (100.0) | 260 (53.4) | 156 (32.0) | 71 (14.6) |  |
| Age (mean (sd)) | 3.99 (1.22) | 3.94 (1.18) | 4.55 (1.08) | 2.94 (0.90) | <0.001 |
| Gender = male (%) | 243 (49.9) | 137 (52.7) | 68 (43.6) | 38 (53.5) | 0.16 |
| Ethnicity = Hispanic (%) | 272 (56.2) | 119 (45.8) | 104 (68.0) | 49 (69.0) | <0.001 |
| Race (%) |  |  |  |  | <0.001 |
| White | 135 (27.7) | 74 (28.5) | 54 (34.6) | 7 (9.9) |  |
| Black | 80 (16.4) | 47 (18.1) | 26 (16.7) | 7 (9.9) |  |
| Asian | 21 (4.3) | 10 (3.8) | 5 (3.2) | 6 (8.5) |  |
| Native Hawaiian/Other Pacific Islander | 31 (6.4) | 31 (11.9) | 0 (0.0) | 0 (0.0) |  |
| American Indian/Alaskan Native | 0 (0.0) | 0 (0.0) | 0 (0.0) | 0 (0.0) |  |
| Other | 220 (45.2) | 98 (37.7) | 71 (45.5) | 51 (71.8) |  |
| Payer source (%) |  |  |  |  | <0.001 |
| Federal Medicaid | 462 (94.9) | 254 (97.7) | 137 (87.8) | 71 (100.0) |  |
| *CHIP | 0 (0.0) | 0 (0.0) | 0 (0.0) | 0 (0.0) |  |
| Employer paid commercial insurance | 1 (0.2) | 1 (0.4) | 0 (0.0) | 0 (0.0) |  |
| Individually paid commercial insurance | 1 (0.2) | 0 (0.0) | 1 (0.6) | 0 (0.0) |  |
| Self-pay | 22 (4.5) | 4 (1.5) | 18 (11.5) | 0 (0.0) |  |
| Other | 1 (0.2) | 1 (0.4) | 0 (0.0) | 0 (0.0) |  |
| **ASA = 2 or 3 (%) | 61 (12.5) | 42 (16.2) | 17 (10.9) | 2 (2.8) | 0.008 |
| †How often has your child had pain in teeth (mean (sd)) | 4.24 (0.99) | 4.03 (1.04) | 4.31 (0.95) | 4.82 (0.52) | <0.001 |
| My child is experiencing pain of the teeth = No (%) | 467 (96.7) | 251 (97.7) | 146 (94.2) | 70 (98.6) | 0.101 |

Means (standard deviations) are presented for continuous variables; frequencies (%) are presented for categorical variables.
GA=General Anesthesia.
\*CHIP = Children's Health Insurance Program.<sup>[28]</sup>
\*\* ASA = American Society of Anesthesiologists Physical Status Classification System.<sup>[29]</sup>
†Variable response options: 5=never, 4=hardly ever, 3=occasionally, 2=often, 1=very often

Significant differences for nearly all variables between patients treated with the three modalities were observed [Table 1]. Compared to children treated with and without general anesthesia, those treated non-surgically were younger in years (mean=2.94, SD=0.90). As well, children treated non-surgically had parental-perceived scores of “pain in their teeth” less often (mean=4.82, SD=0.52) compared to those treated surgically with and without general anesthesia, with the parental-perceived scores categorized as “1=very often,” “2=often,” “3=occasionally,” “4=hardly ever,” and “5=never.”

When compared to the two surgical treatment modalities, Table 2 shows patients treated with non-surgical treatment demonstrated higher, indicating better, average scores for ECOHIS, C-OIDP, and D-CAHPS; and lower average total cost per RVU. Based upon quality, non-surgical treatment had the highest, indicating best, average PRSSQ score (mean=126.13, SD=5.20) compared surgical treatments with GA (mean=115.28, SD=8.22) and without GA (mean=115.85, SD=13.21). More important, significant differences in the value of treatment were observed between the three treatment modalities, with non-surgical treatment having the highest value scores (mean=94.83, SD=72.76) for geometric value and log-transformed value (mean=4.40, SD=0.52).

**Table 2:** Summary of sample comparisons by treatment modalities

|  | Overall | Surgical with GA | Surgical without GA | Non-Surgical | p |
| --- | --- | --- | --- | --- | --- |
| Sum ECOHIS<sup>α</sup> (mean (sd)) | 57.45 (7.61) | 55.59 (7.10) | 58.19 (8.57) | 62.66 (3.25) | <0.001 |
| Average ECOHIS<sup>α</sup> (mean (sd)) | 4.42 (0.58) | 4.28 (0.55) | 4.48 (0.66) | 4.82 (0.25) | <0.001 |
| Sum D-CAHPS<sup>β</sup> (mean (sd)) | 40.30 (4.52) | 40.07 (3.92) | 39.13 (5.24) | 43.68 (3.09) | <0.001 |
| Average D-CAHPS<sup>β</sup> (mean (sd)) | 4.48 (0.50) | 4.45 (0.44) | 4.35 (0.58) | 4.85 (0.34) | <0.001 |
| Sum C-OIDP<sup>χ</sup> (mean (sd)) | 19.30 (2.31) | 19.63 (1.62) | 18.53 (3.32) | 19.79 (1.01) | <0.001 |
| Average C-OIDP<sup>χ</sup> (mean (sd)) | 4.83 (0.58) | 4.91 (0.40) | 4.63 (0.83) | 4.95 (0.25) | <0.001 |
| **Quality** |  |  |  |  |  |
| Total PRSSQ<sup>δ</sup> (mean (sd)) | 117.05 (10.48) | 115.28 (8.22) | 115.85 (13.21) | 126.13 (5.20) | <0.001 |
| ** Average PRSSQ<sup>δ</sup> (mean (sd)) | 4.57 (0.37) | 4.55 (0.28) | 4.49 (0.49) | 4.87 (0.20) | <0.001 |
| **Cost** |  |  |  |  |  |
| Total cost care (mean (sd)) | 381.91 (343.57) | 635.61 (275.75) | 110.80 (99.58) | 48.59 (26.33) | <0.001 |
| RVUs<sup>ε</sup> (mean (sd)) | 29.77 (21.06) | 46.15 (13.02) | 12.49 (11.36) | 7.76 (3.59) | <0.001 |
| † Cost per RVUs<sup>ε</sup> (mean (sd)) | 12.66 (9.60) | 14.23 (6.22) | 12.71 (14.09) | 6.82 (3.68) | <0.001 |
| **Value** |  |  |  |  |  |
| ‡ Value score (mean (sd)) | 58.19 (56.92) | 40.82 (30.08) | 70.45 (70.61) | 94.83 (72.76) | <0.001* |
| Ln-value score (mean (sd)) | 3.79 (0.69) | 3.57 (0.49) | 3.90 (0.84) | 4.40 (0.52) | <0.001 |

\* p < 0.001 by the Kruskal-Wallis test.
\*\* Quality = Average PRSSQ, with higher score indicating higher quality.
GA=General Anesthesia.
† Cost = Cost per RVUs, with lower score indicating lower cost.
‡ Value = Value score = 100 times Quality (\*\*) divided by Cost (†), with higher score indicating higher value.
α ECOHIS: Early Childhood Oral Health Impact Score.<sup>[14]</sup>
β D-CAHPS: Dental Consumer Assessments of Healthcare Providers and Systems. <sup>[15]</sup>
χ C-OIDP: Child-Oral Impacts on Daily Performance.<sup>[16]</sup>
δ PRSSQ: parental-reported symptom and service quality questionnaire.
ε RVUs: Relative Value Units.<sup>[18]</sup>
Means (standard deviations) are presented for continuous variables; frequencies (%) are presented for categorical variables.

After conducting the likelihood ratio test for comparisons of different levels of nesting, two levels of nesting (clinic facility/state) fit the data best, with random effects for states and clinic facilities within each state [Table 3]. After adjusting for patient age, gender, race/ethnicity, and ASA status, the mean log-transformed [confidence interval] value is 0.286 [0.095, 0.477] higher for patients treated with surgical without GA than for those treated with surgical with GA.

**Table 3.** Association of demographic variables and treatment modalities with the log-transformed unweighted value score. Results from linear mixed-effect models with nested random effects (state/facility).

|  | * Two modalities (N=378) |  |  | ** Three modalities (N = 231) |  |  |
| --- | --- | --- | --- | --- | --- | --- |
|  | Estimate | 95% CI | p | Estimate | 95% CI | p |
| Age (continuous) | -0.014 | -0.059, 0.031 | 0.543 | -0.025 | -0.084, 0.034 | 0.399 |
| Gender: female (ref) | - | - | - | - | - | - |
| Gender: male | 0.011 | -0.089, 0.111 | 0.837 | 0.136 | 0.006, 0.266 | 0.039 |
| Ethnicity: non-Hispanic (ref) |  |  |  |  |  |  |
| Ethnicity: Hispanic | 0.062 | -0.076, 0.200 | 0.377 | 0.110 | -0.067, 0.287 | 0.224 |
| ASA 1 (ref) | - | - | - | - | - | - |
| ASA 2 or 3 | -0.186 | -0.343, 0.029 | 0.021 | 0.006 | -0.236, 0.248 | 0.964 |
| Race white (ref) | - | - | - | - | - | - |
| Race Black | 0.108 | -0.075, 0.291 | 0.248 | -0.061 | -0.303, 0.181 | 0.621 |
| Race other | -0.013 | -0.168, 0.142 | 0.865 | 0.022 | -0.216, 0.260 | 0.856 |
| Treatment modality: |  |  |  |  |  |  |
| Surgical with GA (ref) | - | - | - | - | - | - |
| Surgical without GA | 0.286† | 0.095, 0.477 | 0.003 | 0.377‡ | 0.123, 0.631 | 0.004 |
| Non-Surgical | - | - | - | 0.403α | 0.212, 0.594 | <0.001 |

CI: confidence interval; N: subjects with available outcome and covariates.
GA=General Anesthesia.
\* Comparisons between two modalities (surgical with GA vs. surgical with no GA) used data from all states except Hawaii.
\*\*Comparisons between three modalities used data from Maryland and New York only.
† On the original value scale, 0.286 corresponds to 33.1% increase in value for patients treated surgically without GA compared to those treated with GA.
‡ On the original value scale, 0.377 corresponds to 45.7% increase in value for patients treated surgically without GA compared to those treated with GA.
$\alpha$ On the original value scale, 0.403 corresponds to 49.6% increase in value for patients treated non-surgically compared to those treated with GA.

**Table 4.** Distribution of number of subjects by treatment modalities and by state. Patients treated by non-surgical treatment were only observed in Maryland and New York.

| State | Overall | Surgical with GA | Surgical without GA | Non-Surgical |
| --- | --- | --- | --- | --- |
| Arizona | 50 | 16 | 34 | - |
| Hawaii | 35 | 35 | - | - |
| Florida | 54 | 29 | 25 | - |
| Massachusetts | 60 | 27 | 33 | - |
| Maryland | 66 | 29 | 23 | 14 |
| New York | 165 | 85 | 23 | 57 |
| Tennessee | 57 | 39 | 18 | - |
| Total | 487 | 260 | 156 | 71 |

Note:
GA=General Anesthesia.
Only the training locations in Maryland and New York collected data for all three treatment modalities. The non-surgical modality, which includes the use of silver diamine fluoride (SDF) as a caries-arresting agent, was not utilized at the other five training locations because SDF was not commercially available in the United States until 2015 and these five locations had not yet implemented the use of SDF during the data collection period.

Converting the log-transformed value to the original value percent [confidence interval] scale, shows a 33.1% [10.0%, 61.1%] increase in the value score for patients treated surgically without GA compared to those treated with GA. For the comparison on value among three modalities based on the data from Maryland and New York, the mean log-transformed value [confidence interval] of surgical without GA and non-surgical modality are respectively 0.377 [0.123, 0.631] and 0.403 [0.212, 0.594] higher than that of surgical with GA, corresponding to respective increases of 45.7% [13.1%, 87.9%] and 49.6% [23.6%, 81.1%] on the original value scale.

## Discussion

This study confirms its hypothesis that non-surgical treatment yields higher value compared to surgical treatment without GA, which in turn is higher than surgical treatment with GA. The results of this study align with the outcomes of previous studies which independently focus on either oral health related quality of life (**QOL**) or cost outcomes for ECC treatments. To the authors’ knowledge no studies have specifically examined comparative and quantifiable ECC treatment outcomes with a value-based methodology.

Other studies have separately addressed QOL and cost of ECC treatment. There is evidence that the majority of parents perceive higher QOL for their children with ECC after dental treatment under general anesthesia^[21, 22]^ and disease management treatment might result in better quality of care.^[23]^ Additionally, studies have compared costs of ECC treatment and demonstrated the cost for treatment under GA is higher than under sedation, and hospital-based GA treatment has the highest cost.^[24,25]^ Alluding to a value-based model for assessing ECC treatments, a panel of experts at the 2014 Early Childhood Caries Conference concluded disease management treatment may improve outcomes at lower cost and outcomes-based payment models are likely to replace procedure-based payment methods.^[23]^ Another researcher also concluded the U.S. health care system is under pressure to develop a more efficient and cost-effective treatment approach to address early childhood oral health^.[26]^

The primary strength of this study is its quantifiable methodology which integrates quality and cost to extrapolate and demonstrate significant differences in value between three ECC treatment modalities. Although previous studies independently explore QOL and cost for ECC treatment and hint at value-based modeling, this study applies and quantifies a model for comparative value for ECC treatment modalities. Other strengths of this study include the substantial sample size, with post-hoc statistical power at 99.4%, and the broad geographic diversity of the sample population in the U.S.

The limitations of this study include incomplete data, since not all geographic locations collected data for all three treatment modalities; and potential sampling bias, since 94.9% of children were from families of low socioeconomic status (**SES**). Hence, the findings may not be generalizable for the overall U.S. population since low SES populations are at higher risk for chronic health conditions, which may serve as confounding variables.

Future research might address further validation of the PRSSQ survey with a larger sample size and incorporating a pre- and post-treatment survey design; inclusion of total direct cost (not only labor cost as a proxy) to determine if this impacts the cost component of the value-based model; and extension of the data collection period to include more follow-up visits since the Porter-Teisberg model is based upon the full cycle of care and activity-based costing.^[27]^

## Conclusions

Based upon this study’s results, the following conclusions are made:

1. Across the three treatment modalities; non-surgical, surgical without GA, and surgical with GA; there is a significant difference in overall value, cost, and quality.
2. Dentists should consider the value hierarchy when developing informed consent treatment options for the management of ECC; with non-surgical treatment demonstrating higher “value” than surgical without GA treatment, which in turn is higher than surgical with GA treatment.
3. Payers and consumers would benefit by comparing treatment “value,” as available, when choosing institutions and providers for the management of early childhood caries.

## Data Availability

All data produced in the present study are available upon reasonable request to the corresponding author

## Acknowledgments

The authors extend sincere appreciation and acknowledgments to Jay Balzer, DMD, MPH, Dana Cernigliaro, MPH, PhD, Christian Salazar, MPH, PhD, Molly Jung, MPH, PhD, Kera Weiserbs, PhD, MHS; and the former NYU Langone Hospitals-Advanced Education in Pediatric Dentistry residents who supported the study as resident-researchers: Jared Cattron, DMD, Danielle Fernandez, DMD, Shanti Gopalan, DDS, Mehedia Haque, DMD, MPH, Mariam Javaid, DMD, Stephany Liu, DDS, Andrea Lochan, DMD, Jeremy Morris, DDS, Olanrewaju Oye-Somefun, DDS, Andrea Salazar, DDS, Erin Saucier, DMD, Silpa Velivela, DMD, and Pavel Zeylikman, DMD.

## Data availability statement

The data that support the findings of this study are available from the corresponding author upon reasonable request.

